# The Stanford Knee Osteoarthritis PET/MRI Evaluation (SKOPE) Study Protocol

**DOI:** 10.64898/2026.08.26.26361112

**Authors:** Ananya Goyal, Yael Vainberg, Rachel Shalit, Anthony A. Gatti, Feliks Kogan

## Abstract

**Purpose:** The primary objective of the Stanford Knee Osteoarthritis PET/MRI Evaluation (SKOPE) study is to develop and evaluate a multimodal, dynamic [^18^F]NaF PET-MRI framework for characterizing whole-joint physiology and its relationship to osteoarthritis (OA) risk, pain, and disease progression. Specifically, we aim to integrate dynamic PET with quantitative and anatomical MRI, to characterize structural, compositional, and metabolic features across the knee and surrounding musculoskeletal system, evaluate acute tissue responses to exercise, and identify imaging biomarkers associated with OA risk, pain, and disease progression.

**Methods:** The SKOPE study includes multimodal PET-MRI of the knee and surrounding musculoskeletal tissues, with imaging of the knee, tibia, ankle, thigh, hip, pelvis, and lumbosacral spine. Dynamic [^18^F]NaF PET is combined with conventional anatomical MRI and quantitative MRI techniques, including quantitative double-echo steady-state (qDESS) T_2_ mapping of cartilage, Dixon fat-fraction imaging, ultrashort echo time (UTE) T_2_* mapping of short-T_2_ tissues, UTE imaging of tibial bone, and zero echo time (ZTE) imaging for bone morphology and pseudo-CT generation. Additional MRI sequences characterize muscle composition, bone and joint anatomy, intervertebral discs, and regional vascular anatomy. Selected scans are acquired before and after a standardized exercise protocol to assess the acute physiological response of the joint. Automated segmentation is used to generate subject-specific masks of muscles, bones, vertebrae, and intervertebral discs. A subset of the MRI protocol is repeated at 1- and 2-year follow-up to assess longitudinal changes.

**Expected Impact:** By combining dynamic bone metabolic imaging with quantitative measures of cartilage, menisci, muscle, bone, fat, vascular structures, and the spine and hip, the SKOPE protocol provides a whole-joint and multijoint framework for studying the structural, metabolic, and physiological processes associated with OA and pain. Exercise and longitudinal imaging further enable assessment of acute tissue responses and changes over time, supporting the development of quantitative imaging biomarkers for OA risk, pain, and disease progression.

## Introduction

Knee osteoarthritis (OA) is a leading cause of pain and disability and is associated with substantial impairment in physical function and quality of life, as well as considerable societal and economic burden[1,2]. Despite its prevalence, the biological and mechanical processes underlying the initiation and progression of OA remain incompletely understood, and effective disease-modifying treatments remain limited. Importantly, OA is increasingly recognized as a whole-joint disease, involving coordinated alterations across multiple tissues, including cartilage, subchondral bone, menisci, synovium, ligaments, and surrounding musculature[3,4]. A major challenge is therefore the lack of noninvasive imaging approaches that can sensitively characterize early, tissue-specific changes across the joint before established structural abnormalities become apparent.

Radiography remains the primary imaging modality for clinical assessment of knee OA; however, radiographic findings predominantly reflect established morphological changes and have limited sensitivity to early tissue alterations[5,6]. Quantitative magnetic resonance imaging (MRI) provides greater sensitivity to tissue-level changes and can detect compositional and structural abnormalities before conventional radiographic abnormalities become apparent[7–10]. Moreover, quantitative MRI can characterize several components of the joint using tissue-specific techniques. Cartilage T_2_ mapping provides a sensitive measure of cartilage composition[8,11–13], while T_2_*-weighted ultrashort echo time (UTE) imaging enables characterization of tissues with short T_2_* relaxation, including the menisci and ligaments[7,14–16]. Dixon MRI provides quantitative assessment of muscle composition and fat infiltration[17–19]. Together, these approaches provide complementary measures of cartilage, meniscal and ligamentous properties, and muscle composition that extend substantially beyond conventional radiography. However, MRI has limited ability to directly characterize subchondral bone, an important component of the osteochondral unit and a potential early site of pathological change[20].

Positron emission tomography (PET) provides complementary information by enabling noninvasive assessment of tissue physiology and metabolism. In particular, [^18^F]sodium fluoride ([^18^F]NaF) PET is a marker of subchondral bone remodeling, providing information about bone tracer delivery (bone perfusion) and incorporation into the mineralizing bone matrix (bone mineralization)[21,22]. Previous work has demonstrated differences in [^18^F]NaF uptake between individuals with OA and healthy controls[23,24], supporting the potential of [^18^F]NaF PET to detect altered bone physiology associated with OA. Importantly, dynamic PET acquisition and pharmacokinetic modeling can provide information beyond semi-quantitative measures of standardized uptake values (SUVs), including tracer delivery and incorporation[25,26]. Because bone physiology is responsive to mechanical loading, [^18^F]NaF PET also provides an opportunity to assess how bone responds to an acute physiological loading paradigm[27].

Preliminary work from our group demonstrated that physiological joint loading alters [^18^F]NaF uptake in the knee[24,28], providing the basis for using dynamic [^18^F]NaF PET as an imaging-based joint stress test. A standardized stair ascent/descent task provides a reproducible and clinically relevant loading stimulus; therefore, quantifying the acute change in [^18^F]NaF uptake due to stair-climbing may reveal physiological abnormalities that are not apparent under resting conditions.

Combining these complementary measurements within hybrid PET-MRI provides an opportunity to characterize the knee as a whole joint rather than as a collection of isolated tissues[29]. Dynamic [^18^F]NaF PET can characterize bone physiology and its response to loading, while quantitative MRI provides tissue-specific measures of cartilage composition, meniscal and ligamentous properties, and muscle composition. In the present protocol, these measures include cartilage T_2_ mapping using quantitative double-echo steady-state (qDESS) MRI[30], meniscal and ligamentous characterization using ultrashort echo time (UTE) MRI[14], and thigh muscle composition using iterative decomposition of water and fat with echo asymmetry and least-squares estimation-quantitative (IDEAL-IQ) Dixon MRI[19]. Dynamic PET data are evaluated using pharmacokinetic modeling, specifically the Hawkins model[26], together with conventional uptake measures such as SUV. This multimodal framework therefore enables simultaneous assessment of cartilage composition, meniscal and ligamentous properties, and muscle composition, and bone metabolism (both at baseline and in response to a standardized acute loading challenge).

Most previous imaging studies of OA have focused on individuals with established disease and have commonly examined individual tissues, rather than the coordinated physiology of the whole joint. Understanding how abnormalities emerge across the OA continuum therefore requires evaluation of individuals spanning different levels of disease risk and symptom burden. The present protocol includes an asymptomatic cohort in which age, sex, and body mass index (BMI) are evaluated as established OA risk factors, a unilateral knee pain cohort in which painful and contralateral knees can be compared within individuals, and individuals with established OA. This design enables assessment of whether quantitative PET-MRI measures vary across OA risk and disease states and whether regional abnormalities are associated with pain. In particular, within-subject comparison of painful and contralateral knees provides an opportunity to identify spatially concordant abnormalities that may represent potential mechanical pain generators, while the asymptomatic cohort provides a framework for identifying pre-disease alterations associated with OA risk before symptomatic disease is present.

The imaging protocol was subsequently expanded to characterize musculoskeletal changes beyond the knee and to investigate whether observed alterations were localized to the knee or extended across anatomically and mechanically connected regions. For the initial 26 participants, imaging was extended from the tibia through the ankle using a spoiled gradient-echo (SPGR) sequence to support finite element analysis (FEA) of biomechanical strain. Beginning with participant 27, the protocol was changed and expanded to include the hip, lumbar spine, and thigh in addition to the knee. This expanded multijoint acquisition enables exploratory assessment of relationships among bone metabolism, muscle composition, joint structure, biomechanical loading, and pain across multiple anatomical sites, and provides an opportunity to determine whether abnormalities identified by quantitative PET-MRI represent localized joint pathology or broader musculoskeletal alterations.

Finally, the longitudinal component of the study will evaluate whether baseline tissue characteristics and the acute physiological response to loading are associated with subsequent structural and clinical changes over one and two years. An abnormal response to physiological loading may reflect altered joint function that is not captured by static structural measures alone. Longitudinal MRI follow-up therefore provides an opportunity to determine whether baseline dynamic metabolic responses, together with quantitative measures of cartilage, meniscal, ligamentous, and muscle characteristics, are associated with subsequent OA-related tissue changes and pain.

## Study Objective

The primary objective of the Stanford Knee Osteoarthritis PET/MRI Evaluation (SKOPE) study is to develop and evaluate a multimodal, dynamic [^18^F]NaF PET-MRI framework for characterizing whole-joint physiology and its relationship to OA risk, pain, and disease progression. Specifically, we aim to:

1. Determine how whole-joint imaging measures vary across OA risk and symptom states, including age, sex, and BMI in asymptomatic individuals, unilateral knee pain, and established OA, and identify regional imaging abnormalities that may represent potential mechanical pain generators.
2. Evaluate the longitudinal relevance of these imaging biomarkers by determining whether baseline tissue characteristics and dynamic responses to loading are associated with subsequent structural and clinical changes.
3. Evaluate the physiological response of the knee to acute mechanical loading using pre- and post-exercise [^18^F]NaF PET, and determine whether loading-induced changes provide information beyond conventional static imaging measures.
4. Explore the relationship between knee abnormalities and broader musculoskeletal physiology through imaging of the hip, lumbar spine, and thigh, determining whether alterations are localized to the knee or extend across anatomically connected regions.

## Materials and Methods

### 1. Participants and Cohorts

Participants aged 18–80 years were recruited from the community between February 2023 and December 2024 through advertisements, including flyers, online postings, and institutional mailing lists. Interested individuals completed an initial online screening questionnaire followed by a telephone interview to assess eligibility based on age, knee symptoms, prior knee injury or surgery, and medical history. Eligible participants were enrolled after providing written informed consent under an Institutional Review Board-approved, HIPAA-compliant protocol. Individuals with contraindications to MRI, including incompatible metal implants or claustrophobia, were excluded.

Participants were recruited into three cohorts based on their clinical status:

#### Asymptomatic cohort

Healthy adults without a history of knee pain, knee injury, or prior knee surgery were recruited to investigate associations between OA risk factors and subclinical whole-joint imaging measures. Seventy-one asymptomatic adults aged 20–80 years (36 female) were enrolled. Recruitment targeted approximately balanced representation of males and females across age decades, with approximately five males and five females recruited per decade from 20–70 years; recruitment of participants aged 70–80 years was lower because of limited availability of eligible participants. Age, sex, and anthropometric measures were recorded. Participants with contraindications to PET or MRI, including incompatible metal implants, as well as systemic musculoskeletal or metabolic conditions that could affect joint tissues, were excluded.

#### Unilateral knee pain cohort

Twenty-two participants with unilateral knee pain were recruited to investigate potential tissue-specific and regional imaging correlates of pain. Participants were required to have unilateral knee symptoms and were excluded for contraindications to PET or MRI, including incompatible metal implants, as well as medical conditions that could substantially affect musculoskeletal tissues. Participants with bilateral or systemic musculoskeletal conditions were excluded when these conditions prevented identification of a clinically distinct painful and contralateral knee.

#### Knee OA cohort

Forty participants aged 30–80 years with self-reported, symptomatic, and clinically diagnosed knee OA were recruited to investigate whole-joint imaging abnormalities associated with established symptomatic OA. Participants with either unilateral or bilateral knee OA were included. Participants were excluded for contraindications to PET or MRI, including incompatible metal implants, as well as systemic musculoskeletal or metabolic conditions that could affect joint tissues.

Participants in the knee pain and knee OA cohorts were age- and sex-matched to participants in the asymptomatic cohort, with matching performed within ±3 years of age. This design enabled comparisons across asymptomatic individuals with varying levels of OA risk, symptomatic individuals with unilateral knee pain, and participants with established symptomatic OA while accounting for major demographic differences between cohorts.

All participants in the knee pain and knee OA cohorts were invited to return for one- and two-year follow-up MRI scans. For the asymptomatic cohort, only participants over the age of 50 years were invited to return for the one-year follow-up MRI scan; all asymptomatic cohort participants were invited to return for the two-year follow-up MRI.

### 2. PET-MRI Image Acquisition

All participants underwent bilateral lower-extremity imaging on a 3T GE Signa PET/MRI scanner (GE Healthcare, Waukesha, WI, USA). Bilateral knee imaging was performed using two medium-sized flexible 16-channel receive-only phased-array coils (Neocoil, Milwaukee, WI, USA), while imaging of the hip, lumbar spine, thigh, and other lower-extremity regions was performed using the integrated 14-channel posterior spine table phased-array coil. Imaging was performed before and immediately after a standardized acute mechanical loading task consisting of ascending and descending eight flights of stairs (135 steps up and 135 steps down). The dynamic PET acquisition protocol was maintained across participants and imaging time points, whereas the MRI protocol varied according to the imaging time point and study objectives to provide complementary structural, compositional, and quantitative information across multiple tissues.

#### 2.1 PET Imaging Protocol

Participants received an intravenous hand injection of 92.5 MBq (approximately 2.5 mCi) of [^18^F]sodium fluoride ([^18^F]NaF) through an antecubital venous catheter. Simultaneous PET/MRI acquisition was initiated just before the start time of tracer administration, and dynamic PET list-mode data were acquired continuously for 30 minutes over the bilateral knees[31]. A two-point Dixon MRI sequence was acquired during the PET examination for MR-based attenuation correction.

Following the baseline acquisition, participants completed an acute mechanical loading task consisting of ascending and descending eight flights of stairs (135 steps ascending and 135 steps descending). Immediately following exercise, participants underwent a second PET/MRI examination using the same PET acquisition protocol, including a second 92.5-MBq [^18^F]NaF injection, to quantify exercise-induced changes in tracer delivery and uptake.

For the post-exercise examination, a 3-minute dynamic PET acquisition was performed immediately before the second tracer injection to characterize residual tracer activity from the baseline tracer administration. Following the second injection, dynamic PET data were again acquired continuously for 30 minutes over the bilateral knees.

##### Lower-leg cohort

For participants 1–26, the PET acquisition was additionally extended to the distal lower extremity after 30 minutes of bilateral knee imaging. Approximately 4 minutes of PET acquisition were performed at each of three additional bed positions encompassing the ankle, distal tibia/shin, and proximal tibia immediately inferior to the knee. These additional acquisitions were performed at both baseline and following exercise.

##### Spine-to-knee cohort

Beginning with participant 27, the protocol was further expanded to include additional PET bed positions encompassing the lumbar spine, hip, and thigh, with approximately 4 minutes of acquisition at each location at both baseline and post-exercise, followed by 4 minutes of additional bilateral knee acquisition. This multibed acquisition enabled exploratory assessment of tracer uptake across anatomically and mechanically connected regions.

Dynamic PET data from both baseline and post-exercise examinations were reconstructed into multiple image series for image-derived arterial input function (AIF) estimation, regional time-activity curve (TAC) analysis, PET angiography (PETA), and static end-of-scan quantification. All PET reconstructions were performed using time-of-flight ordered-subset expectation maximization (TOF-OSEM; three iterations), with a β-value of 350, with corrections for radioactive decay, attenuation, scatter, random coincidences, and detector dead time. Reconstructed PET images had a voxel size of 1.3 x 1.3 x 2.78 mm^3^.

For image-derived AIF estimation, the dynamic acquisition was reconstructed using frames of 2 x 5 seconds, 20 x 1 seconds, 10 × 10 seconds, 10 x 30 seconds, 5 x 1 minutes, and 9 x 2 minutes to capture the rapid tracer bolus and subsequent blood clearance. Regional bone TACs were reconstructed using 6 × 10 seconds, 10 x 1 minutes, and 9 x 2 minutes frames. PET angiography images were reconstructed from the first 15 seconds following tracer bolus arrival to visualize the arterial vasculature. Static end-of-scan images were reconstructed from the final 5 minutes of the dynamic acquisition (25–30 minutes post-injection) for quantitative assessment of tracer uptake.

**Figure 1.**
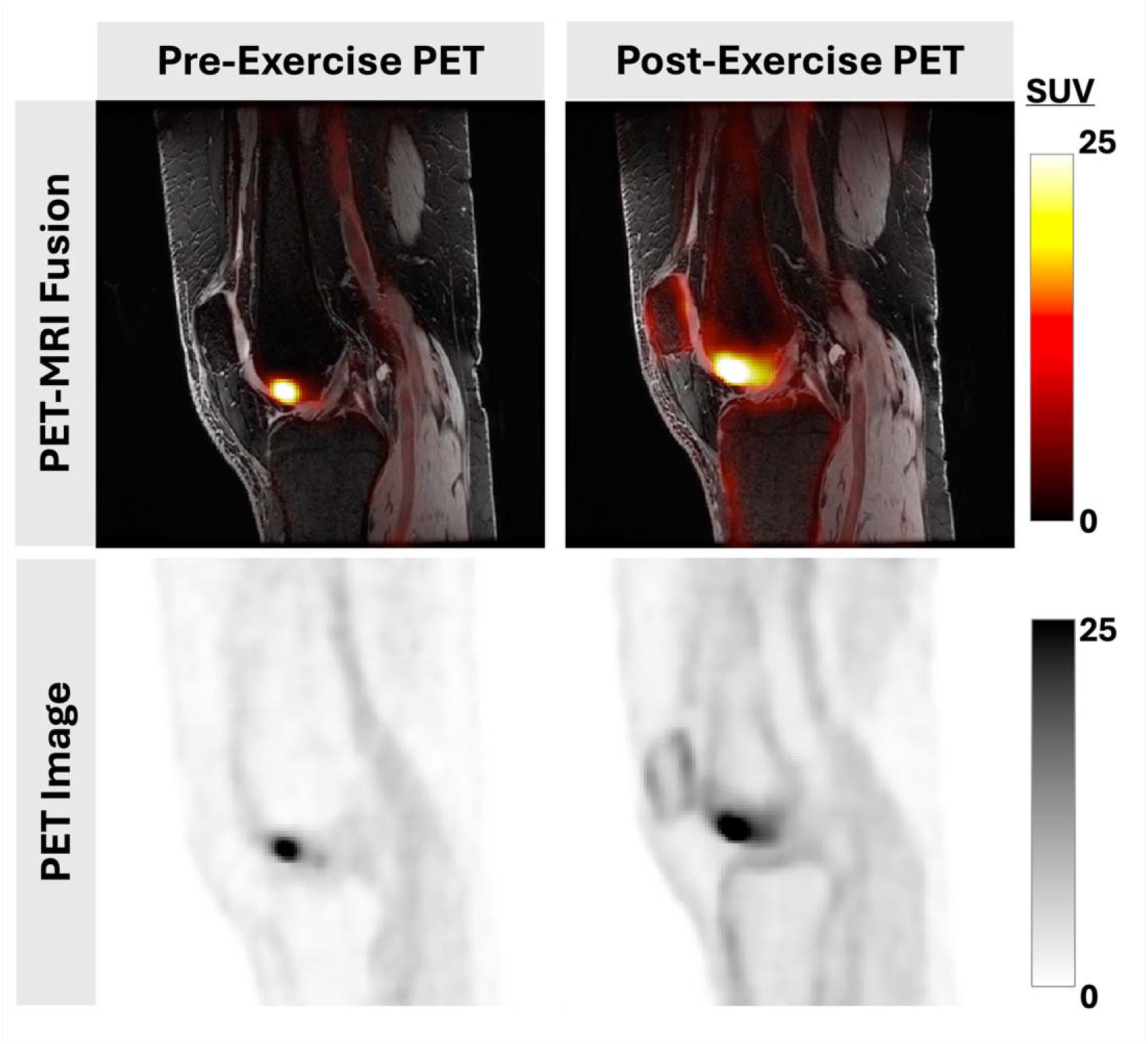
Representative PET-MRI images before and after exercise. Representative fused PET-MRI images acquired before and after the exercise protocol, along with original PET images; PET signal hotspots represent areas of potentially increased bone perfusion / bone mineralization.

#### 2.2 MR Imaging Protocol

MRI acquisitions were performed concurrently with PET and were selected to provide complementary information on joint structure, tissue composition, and tissue-specific quantitative properties. The MRI protocol differed between baseline and post-exercise examinations and evolved over the course of the study to accommodate additional tissue-specific and multijoint measurements. A quantitative double-echo steady-state (qDESS) sequence was acquired at both time points for bilateral knee tissue segmentation and cartilage T_2_ relaxometry.

For the ***lower-leg cohort***, in participants 1–26, the MRI protocol was extended to the lowe included imaging extending from the knee through the tibia and ankle. In particular, an SPGR acquisition was obtained to support subsequent finite element analysis (FEA) of biomechanical strain.

For the ***spine-to-knee cohort***, participant 27 onwards, the MRI protocol was expanded to include the hip, lumbar spine, and thigh, enabling characterization of additional musculoskeletal tissues and regions within the same PET-MRI examination.

The complete MRI acquisition parameters, including sequence-specific acquisition parameters and the distribution of sequences across baseline and post-exercise examinations, are provided below.

##### Knee MRI

###### Anatomical MRI Scans

All acquisition parameters for anatomical MRI scans are provided in **Table 1**, and representative images are shown in **Figure 2**.

**Figure 2.**
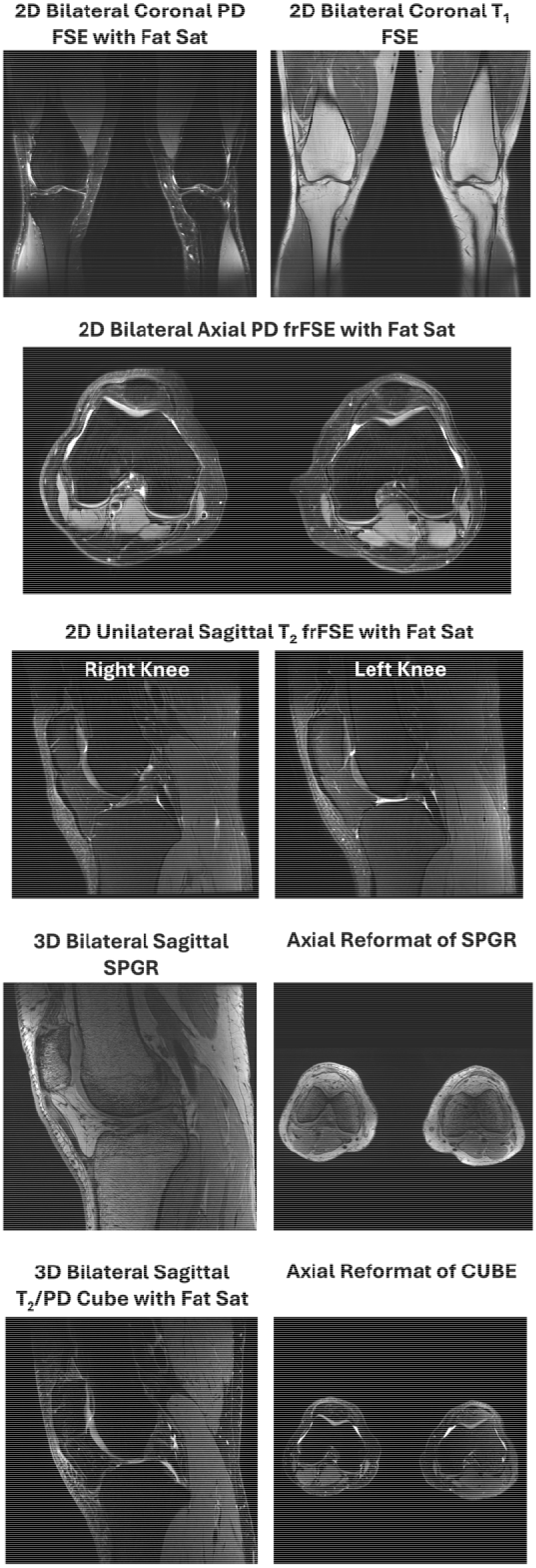
Representative images from the anatomical MRI scans, including multiple 2D FSE and 3D sequences. Together, these sequences provide comprehensive visualization of knee joint anatomy, including cartilage, menisci, ligaments, bone marrow, joint fluid, periarticular soft tissues, and subcutaneous and intra-articular fat.

**Table 1.** Acquisition parameters for the conventional anatomical MRI sequences acquired for knee assessment.

| Scan Parameter | Sagittal T<sub>2</sub> frFSE FS | Coronal PD FSE FS | Axial PD frFSE FS | Coronal T<sub>1</sub> FSE | Sagittal T<sub>2</sub>/PD Cube FS | Sagittal SPGR |
| --- | --- | --- | --- | --- | --- | --- |
| 2D/ 3D | 2D | 2D | 2D | 2D | 3D | 3D |
| Uni/ Bi | Uni | Bi | Bi | Bi | Bi | Bi |
| TR (ms) | 6938 | 9383 | 5463 | 843 | 1000 | 10 |
| TE (ms) | 63 | 24 | 24 | 9 | 64 | 5 |
| FA (°) | 111 | 111 | 111 | 111 | 90 | 20 |
| ETL | 18 | 8 | 8 | 3 | 50 | 1 |
| # Slices | 60 | 65 | 36 | 32 | 320 | 240 |
| Slice thickness (mm) | 1.5 | 1.5 | 3 | 3 | 1 | 1.5 |
| Acq matrix | 352 x 256 | 640 x 512 | 640 x 640 | 768 x 640 | 320 x 320 | 420 x 384 |
| Recon matrix | 512 x 512 | 1024 x 1024 | 1024 x 1024 | 1024 x 1024 | 512 x 512 | 512 x 512 |
| FOV (cm) | 16 x 16 | 34 x 34 | 34 x 34 | 32 x 32 | 16 x 16 | 16 x 16 |
| Recon in-plane resolution (mm<sup>2</sup>) | 0.31 x 0.31 | 0.33 x 0.33 | 0.33 x 0.33 | 0.31 x 0.31 | 0.31 x 0.31 | 0.31 x 0.31 |
| Acc (phase x slice) | 2 x 1 | 3 x 1 | 2 x 1 | 3 x 1 | 2 x 3 | 2 x 1 |
| NEX | 1 | 1 | 1 | 1 | 1 | 1 |
| BW (Hz/pixel) | 163 | 81 | 81 | 163 | 325 | 122 |
| TA (min:sec) | 3:01 | 3:91 | 2:46 | 2:20 | 4:08 | 3:43 |

qDESS quantitative double-echo steady-state; SPGR spoiled gradient recalled echo; UTE ultrashort echo time; ZTE zero echo time; Uni unilateral; Bi bilateral; TR repetition time; TE echo time; FA flip angle; ETL echo train length; Acq acquired; Recon reconstructed; FOV field of view; Acc acceleration factor; NEX number of excitations; BW bandwidth; TA acquisition time.

###### 1. Unilateral sagittal T_2_-weighted fast recovery fast spin echo (frFSE) with fat saturation (FS)

A unilateral sagittal two-dimensional (2D) T_2_-weighted frFSE sequence with fat saturation was acquired for evaluation of fluid-sensitive abnormalities and internal derangements of the knee (one scan per knee). This sequence provides high sensitivity to joint fluid, making it particularly useful for detection of joint effusion, synovitis, bone marrow lesions/edema-like signal, cartilage abnormalities, meniscal and ligamentous pathology, and other soft-tissue injuries.

###### 2. Bilateral coronal proton density (PD)-weighted fast spin echo (FSE) with FS

A bilateral coronal 2D PD-weighted FSE sequence with fat saturation was acquired to provide comprehensive assessment of the knee joint and periarticular soft tissues. The sequence is well suited for evaluation of meniscal morphology and tears, collateral ligaments, cartilage defects, bone marrow lesions, joint fluid, and other structural abnormalities, while enabling efficient bilateral comparison.

###### 3. Bilateral axial PD-weighted frFSE with FS

A bilateral axial 2D PD-weighted frFSE sequence with fat saturation was acquired for assessment of structures best visualized in the axial plane. This sequence provides high-quality evaluation of patellofemoral cartilage, patellar alignment, quadriceps and patellar tendons, Hoffa’s fat pad, synovium, joint fluid, and periarticular soft-tissue abnormalities.

###### 4. Bilateral coronal T_1_-weighted FSE

A bilateral coronal 2D T_1_-weighted FSE sequence was acquired for anatomical delineation and assessment of bone marrow and soft-tissue composition. T_1_-weighted imaging provides strong anatomical contrast and is particularly useful for evaluating bone marrow abnormalities, subcutaneous and periarticular fat, and overall osseous morphology.

###### 5. Bilateral sagittal T_2_-weighted/ PD-weighted Cube with FS

A bilateral sagittal three-dimensional (3D) Cube sequence with T_2_-weighted/PD-weighted contrast and fat saturation was acquired to provide high-resolution anatomical imaging of the knee. The sequence enables multiplanar reformats and assessment of cartilage morphology, menisci, cruciate ligaments, bone marrow, joint fluid, and other intra-articular structures, with particular utility for detecting subtle cartilage defects, meniscal abnormalities, ligamentous pathology, and bone marrow lesions.

###### 6. Bilateral sagittal spoiled gradient-recalled echo (SPGR)

A bilateral sagittal 3D SPGR sequence was acquired to provide high-resolution morphological assessment of the knee, with particular utility for cartilage visualization. The sequence provides strong cartilage-to-fluid contrast and high spatial resolution, facilitating assessment of cartilage thickness, surface morphology, focal defects, and other structural abnormalities, as well as detailed evaluation of overall joint anatomy.

###### 7. Bilateral sagittal quantitative double-echo steady-state (qDESS)

A three-dimensional (3D) sagittal bilateral quantitative double-echo steady-state (qDESS) sequence was acquired for quantitative T_2_ relaxometry and automated segmentation of knee tissues, including cartilage, bone, and menisci. The sequence provided high-resolution coverage of both knees to enable simultaneous assessment of tissue morphology and quantitative cartilage composition, while enabling multiplanar reformats. qDESS scans were acquired twice– both before and after the exercise protocol. Acquisition parameters are provided in **Table 2**, and representative images are shown in **Figure 3**.

**Figure 3.**
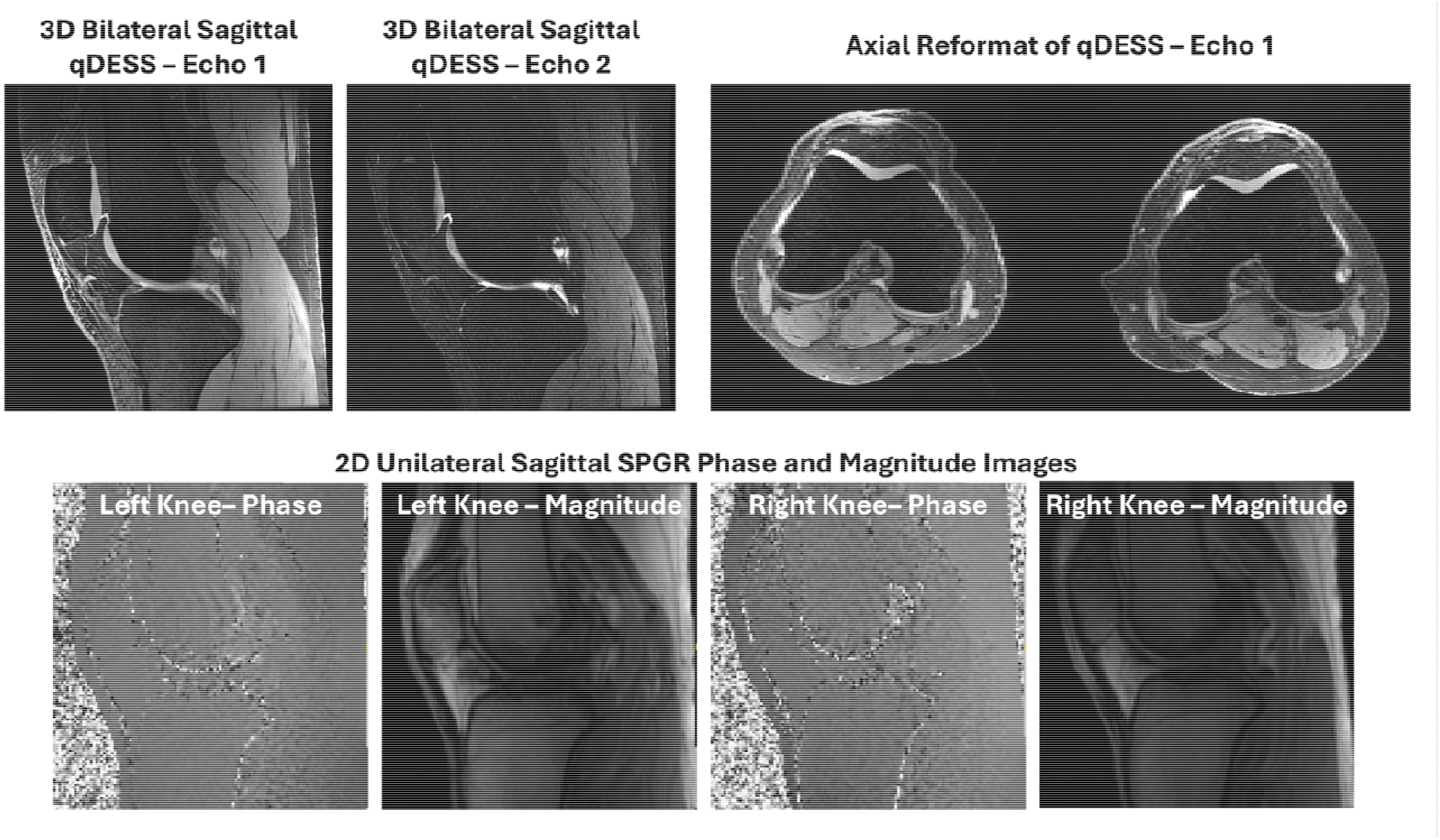
Representative bilateral sagittal qDESS images, together with sagittal SPGR images for B_1_ correction. qDESS images demonstrate high-resolution bilateral knee anatomy and cartilage visualization, while the phase and magnitude SPGR images can be used for B_1_ correction, to characterize spatial variations in transmit field used for voxel-wise correction of qDESS-derived T_2_ measurements.

**Table 2.**
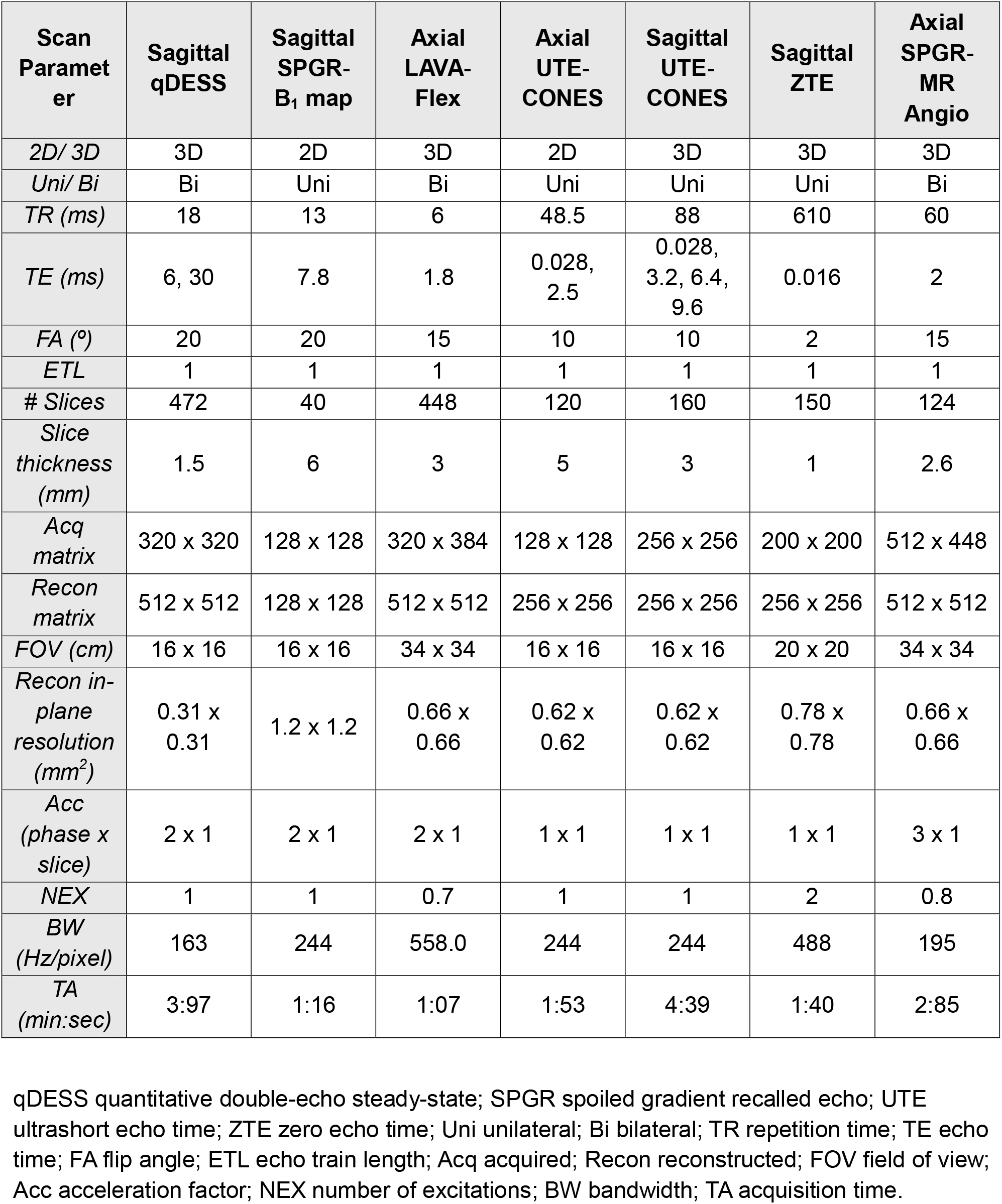
Acquisition parameters for the quantitative MRI and MR angiography sequences.

###### 8. Unilateral sagittal SPGR for B_1_ correction

Two unilateral sagittal 2D SPGR scans (one per leg) were acquired to characterize spatial variations in transmit field across the knee and enable correction of B_1_-related signal inhomogeneities in quantitative MRI measurements. The resulting B_1_ correction maps can be coregistered to the qDESS images and used for voxel-wise correction of the measured signal prior to T_2_ relaxometry. Scans for B_1_ correction were acquired twice– both before and after the exercise protocol. Acquisition parameters are provided in **Table 2**, and representative images are shown in **Figure 3**.

###### 9. Bilateral axial LAVA Flex

A bilateral axial 3D LAVA Flex sequence was acquired for quantitative assessment of tissue fat composition across the knee and surrounding soft tissues. The sequence provides water- and fat-separated images that enable calculation of fat fraction. Fat fraction maps can be used to characterize intramuscular fat infiltration across the thigh and periarticular muscles, as well as fat composition within the infrapatellar (Hoffa’s) fat pad and other regional adipose tissues. Acquisition parameters are provided in **Table 2**, and representative images are shown in **Figure 4**.

**Figure 4.**
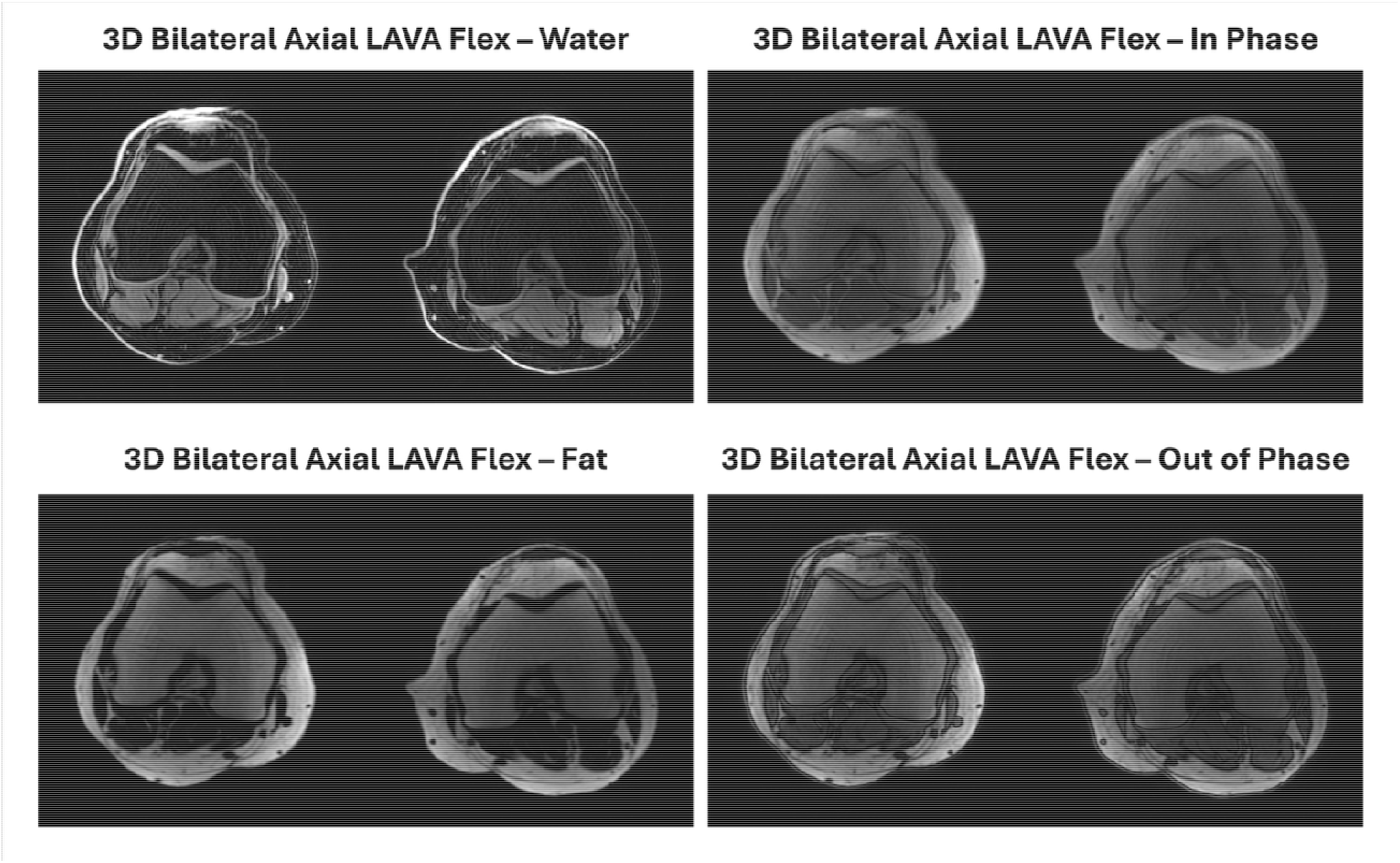
Representative bilateral axial LAVA Flex water- and fat-separated images (as well as in-phase and out-of-phase images). Fat fraction maps (derived as the ratio of fat to fat + water) can enable quantitative assessment of fat composition across the thigh and periarticular musculature, as well as regional adipose tissues including Hoffa’s fat pad.

###### 10. Unilateral axial ultrasort echo time (UTE) of tibial long bone

A unilateral axial 3D UTE CONES sequence was acquired to characterize short-T_2_ tissues and assess changes in bone following exercise in the tibial long bone (one scan per knee). The dual-echo UTE acquisition provides sensitivity to rapidly decaying signal components and can be used to derive a UTE signal-based porosity index from the ratio of the first and second echoes (0.028 ms and 2.5 ms). Repeat acquisitions performed before and after exercise may also enable assessment of exercise-induced changes in tibial long bone signal, providing a potential imaging marker of transient alterations in bone perfusion. Acquisition parameters are provided in **Table 2**, and representative images are shown in **Figure 5**.

**Figure 5.**
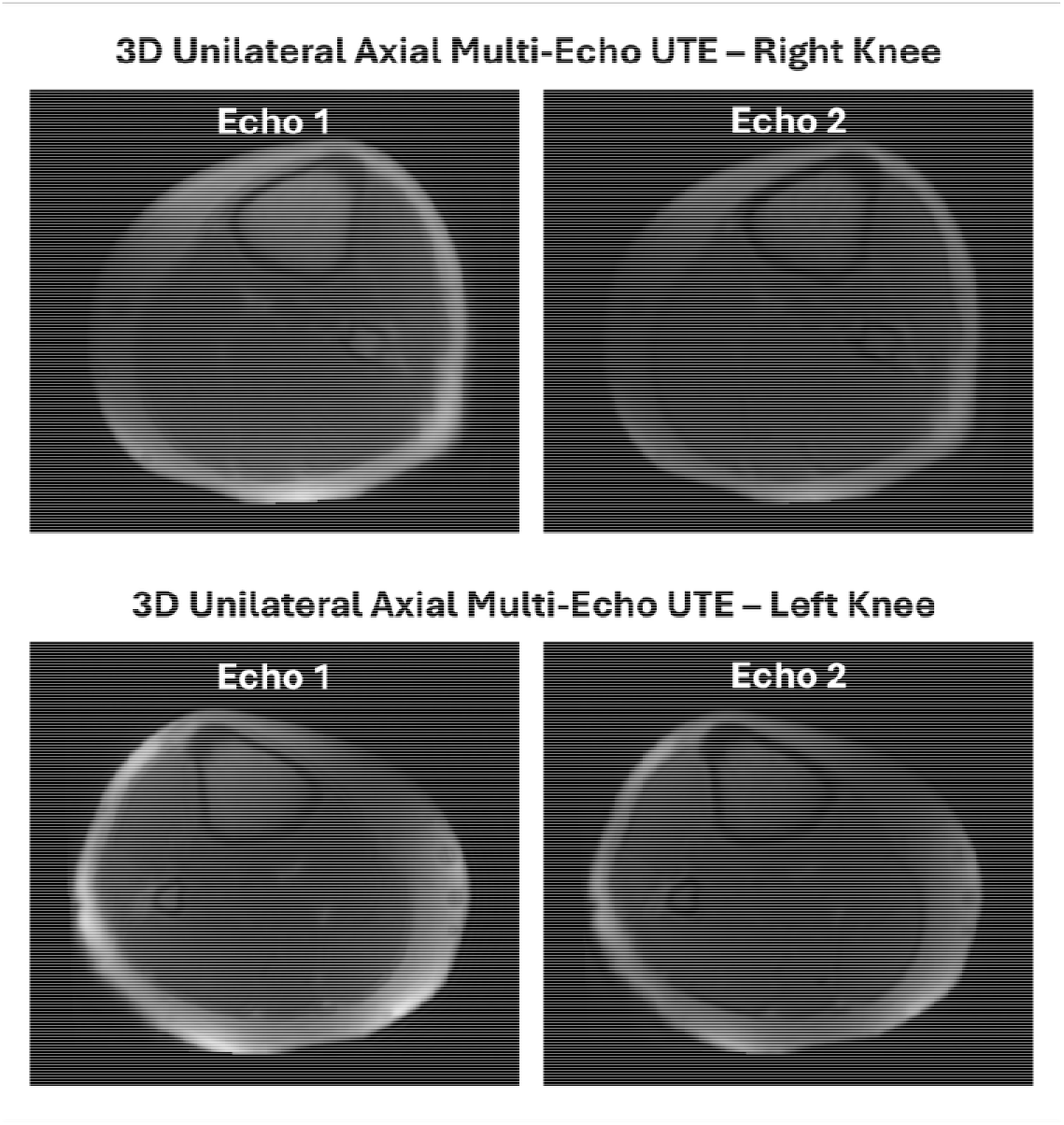
Representative axial dual-echo UTE CONES images of the tibial long bone, acquired unilaterally for each leg. Images demonstrate the short-T_2_-sensitive UTE signal within the tibial cortex and other osseous structures; the ratio of the second echo to the first echo can provide a UTE-based porosity index, which can potentially examine exercise-related changes in tibial bone perfusion.

###### 11. Unilateral sagittal UTE of the knee

A unilateral sagittal 3D UTE CONES sequence of the knee was acquired for quantitative characterization of tissues with short T_2_ relaxation times, including the menisci, tendons, and ligaments (one scan per knee). The multi-echo acquisition (four echoes) can enable generation of quantitative mono-exponential T_2_* maps, providing measures of tissue composition and microstructural organization that are not readily captured with conventional MRI. T_2_* mapping can be used to characterize the menisci and major knee ligaments and tendons, with sensitivity to tissue degeneration and structural alterations associated with joint pathology. Acquisition parameters are provided in **Table 2**, and representative images are shown in **Figure 6**.

**Figure 6.**
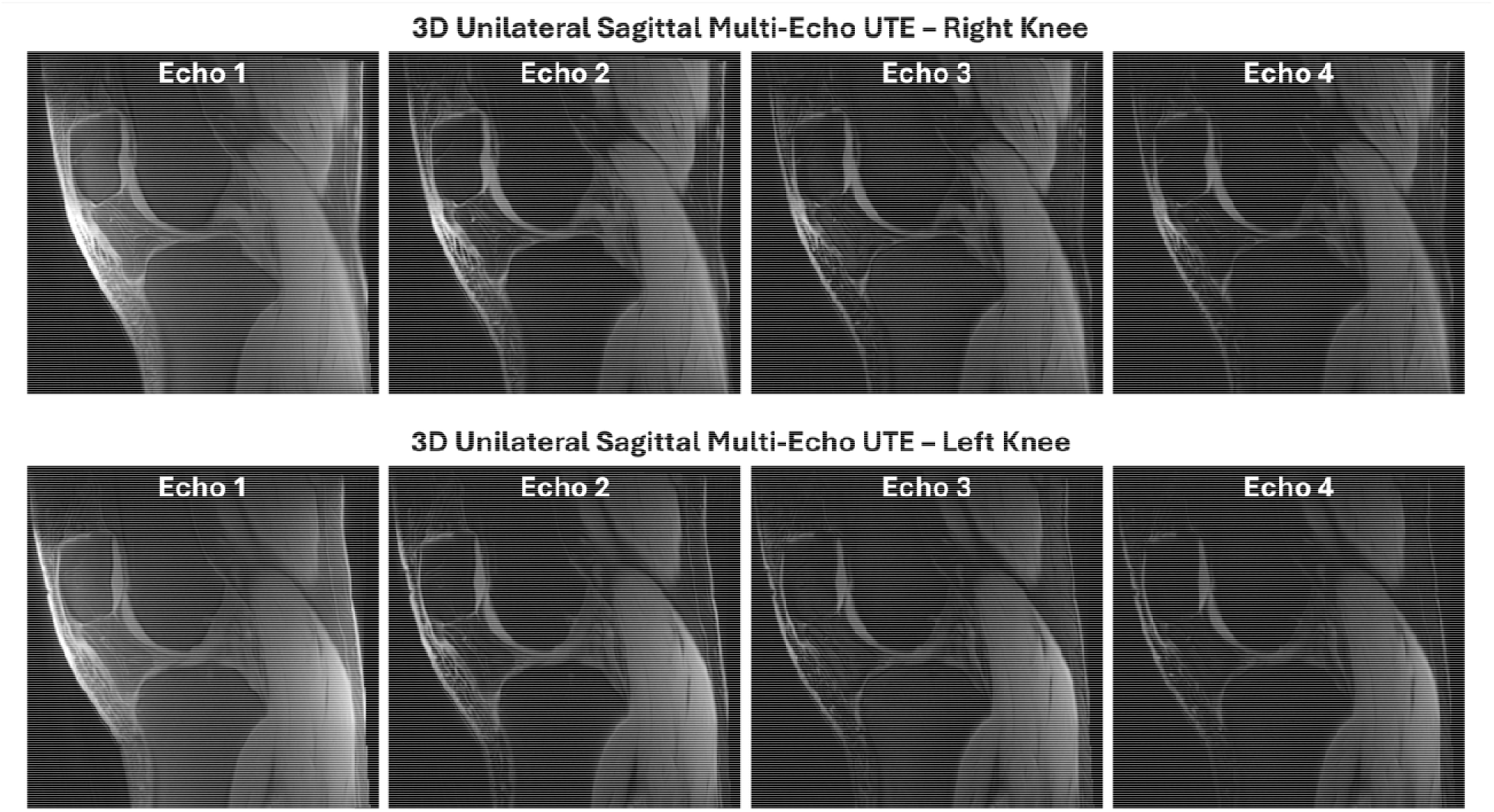
Representative sagittal multi-echo UTE CONES images, acquired unilaterally, that can be used for mono-exponential T_2_* mapping for the menisci, tendons, and ligaments, providing quantitative characterization of short-T_2_ tissues and their underlying composition and microstructural organization.

###### 12. Unilateral sagittal zero echo time (ZTE)

A unilateral sagittal 3D ZTE sequence was acquired for high-resolution characterization of bone morphology (one scan per knee). The high spatial resolution and sensitivity to cortical bone can allow detailed three-dimensional modeling of bone shape and joint geometry. ZTE images can additionally be used to generate synthetic or pseudo-computed tomography (pseudo-CT) images, providing CT-like representation of bone density and cortical structure without ionizing radiation. Acquisition parameters are provided in **Table 2**, and representative images are shown in **Figure 7**.

**Figure 7.**
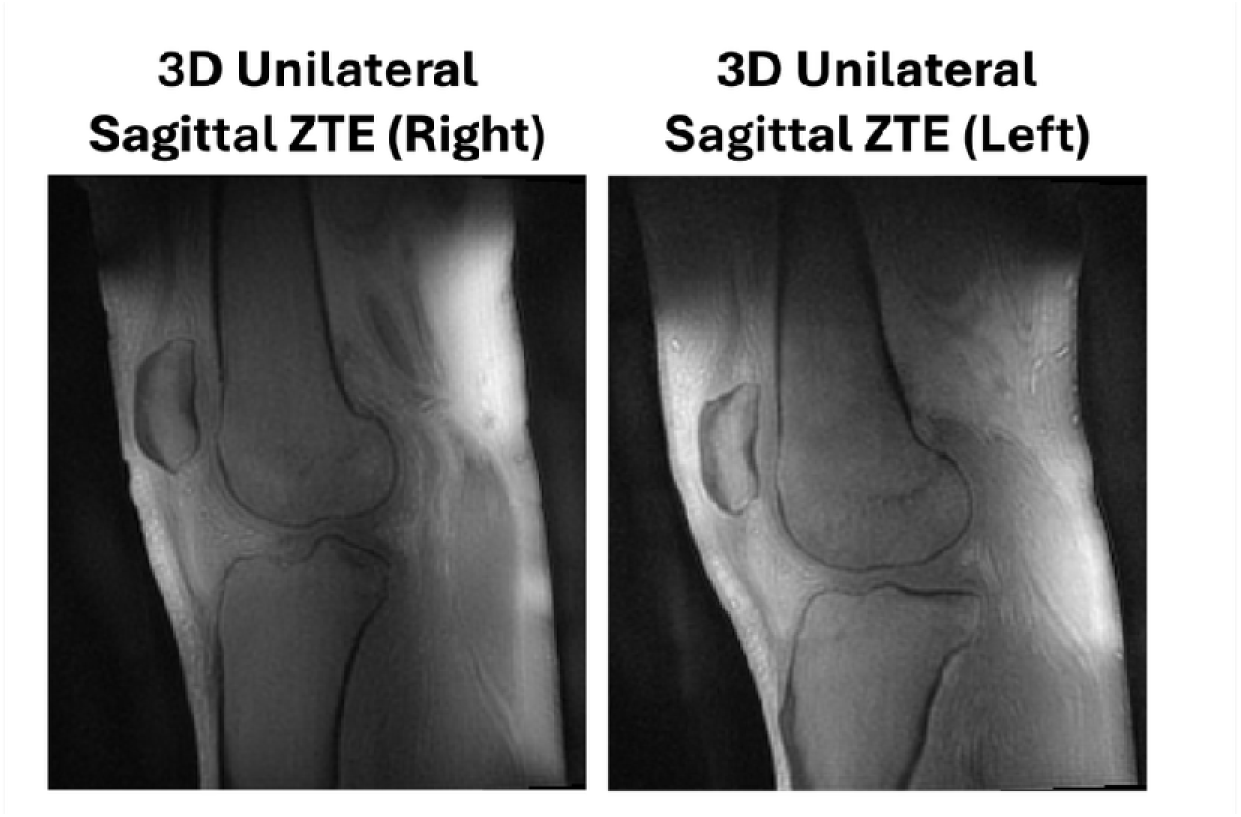
Representative sagittal ZTE images of the knee demonstrating high-resolution visualization of osseous anatomy and cortical bone. ZTE-derived images can be used for 3D bone representations and/or synthetic pseudo-CT images for visualization of bone morphology

###### 13. Bilateral axial SPGR for MR angiography

A bilateral axial 3D vascular time-of-flight (ToF) SPGR sequence was acquired for MR angiography to visualize the popliteal artery of both legs. The sequence provided high-resolution coverage of the bilateral knees and surrounding soft tissues, enabling assessment of popliteal artery anatomy. Acquisition parameters are provided in **Table 2**, and representative images are shown in **Figure 8**.

**Figure 8.**
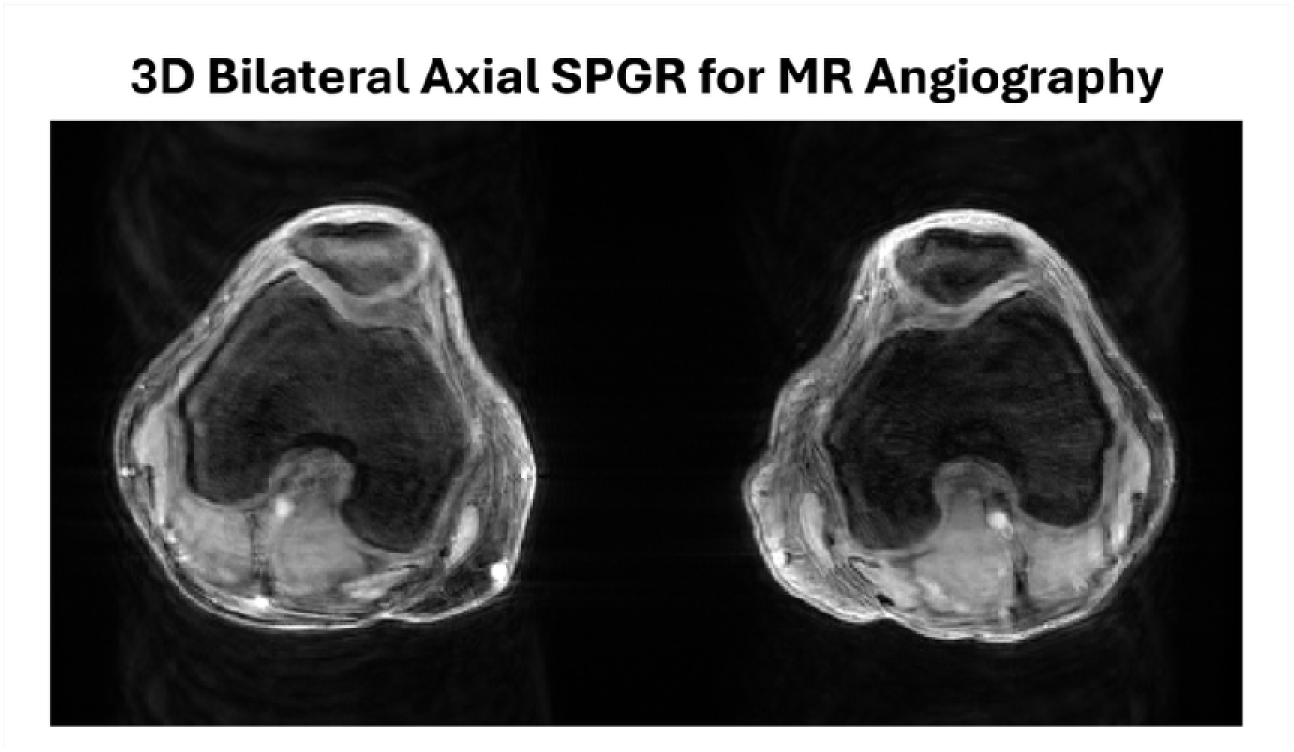
Representative bilateral axial time-of-flight (ToF) SPGR MR angiography images demonstrating visualization of the popliteal arteries at the level of the knee.

##### Lower-Leg Protocol

###### 1. Bilateral axial SPGR

A bilateral axial 3D SPGR sequence was acquired to provide high-resolution anatomical visualization of the distal tibia, fibula, and ankle joint. The sequence was used to characterize bone morphology and overall ankle anatomy, including the tibial tuberosity, tibial longbone, and distal tibia, and to support assessment of bone strain using finite element analysis (FEA). The same scans were repeated before and after exercise. Acquisition parameters are provided in **Table 3**, and representative images are shown in **Figure 9**.

**Figure 9.**
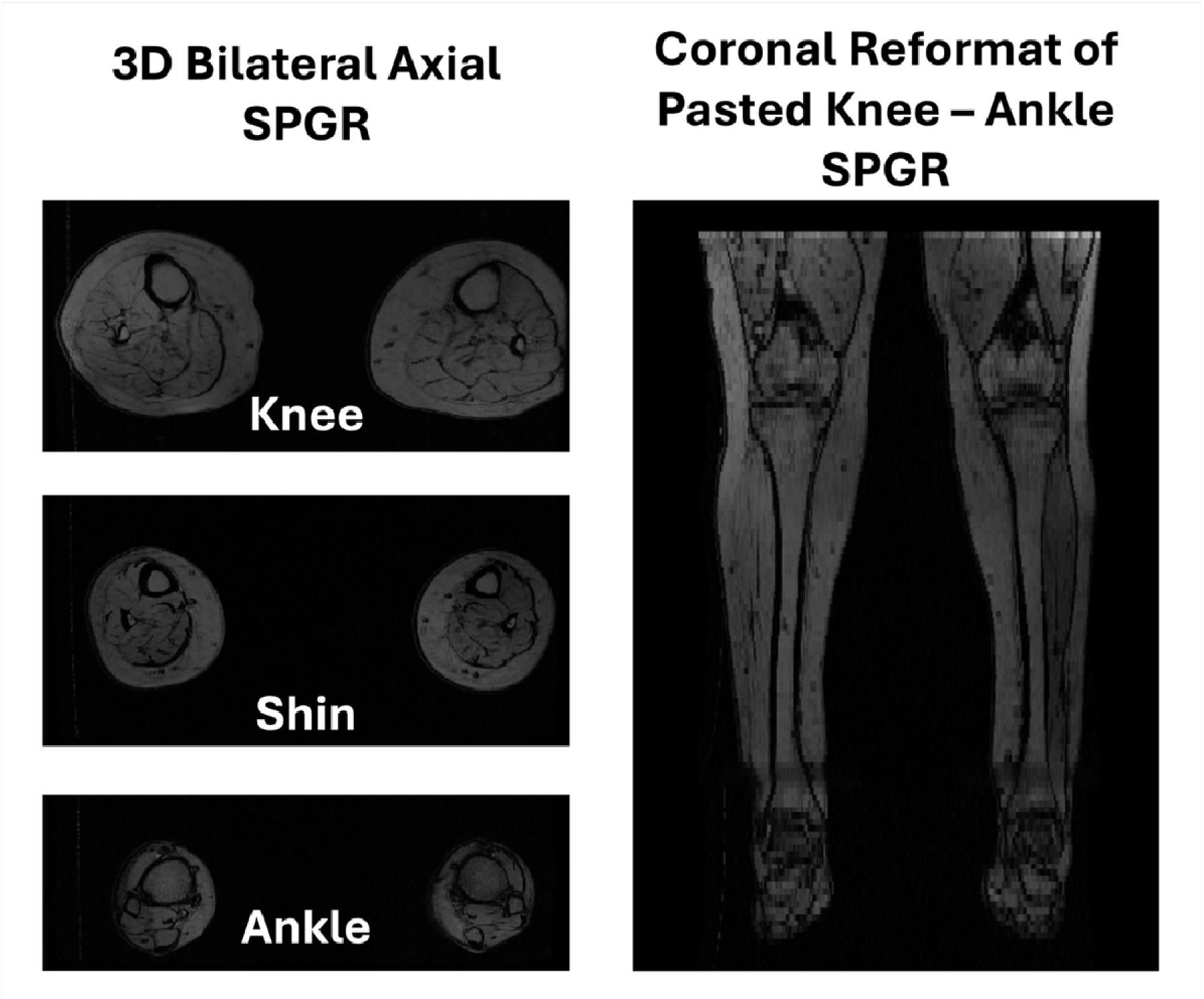
Representative bilateral axial 3D SPGR images of the lower leg and ankle. Images show high-resolution visualization of the tibial tuberosity, tibial shaft, distal tibia, fibula, and ankle joint anatomy. These images provide anatomical input for assessment of bone geometry and exercise-related changes in bone strain using finite element analysis (FEA).

**Table 3.** Acquisition parameters for MRI of joints beyond the knee, specifically the shin and ankle (lower-leg cohort), and lumbosacral spine, hip, and thigh (spine-to-knee cohort).

| Scan Parameter | Knee, Shin, Ankle | Lumbosacral Spine |  | Hip |  | Thigh |  |
| --- | --- | --- | --- | --- | --- | --- | --- |
|  | Axial SPGR | Axial IDEAL-IQ | Sagittal T<sub>2</sub> Cube FS | Coronal PD frFSE FS | Axial T<sub>2</sub> Cube FS | Axial IDEAL-IQ | Axial SPGR |
| *2D/ 3D* | 3D | 3D | 3D | 2D | 3D | 3D | 3D |
| *Uni/ Bi* | Bi | - | - | Bi | Bi | Bi | Bi |
| *TR (ms)* | 12 | 8 | 1500 | 3868 | 1500 | 9 | 12 |
| *TE (ms)* | 5.8 | 1, 2, 3, 4, 5, 6 | 59 | 30 | 59 | 1, 2, 3, 4, 5, 6 | 5.8 |
| *FA (°)* | 10 | 4 | 90 | 111 | 90 | 4 | 10 |
| *ETL* | 1 | 3 | 80 | 8 | 80 | 3 | 1 |
| *# Slices* | 52 | 240 | 100 | 30 | 160 | 240 | 54 |
| *Slice thickness (mm)* | 5 | 8 | 1.97 | 3 | 2 | 8 | 8 |
| *Acq matrix* | 640 x 640 | 290 x 350 | 500 x 420 | 640 x 420 | 320 x 320 | 274 x 360 | 640 x 640 |
| *Recon matrix* | 1024 x 1024 | 512 x 512 | 512 x 512 | 1024 x 1024 | 512 x 512 | 512 x 512 | 1024 x 1024 |
| *FOV (cm)* | 32 x 32 | 46 x 46 | 30 x 30 | 40 x 40 | 36 x 36 | 40 x 40 | 32 x 32 |
| *Recon in-plane resolution (mm<sup>2</sup>)* | 0.31 x 0.31 | 0.89 x 0.89 | 0.59 x 0.59 | 0.39 x 0.39 | 0.70 x 0.70 | 0.78 x 0.78 | 0.31 x 0.31 |
| *Acc (phase x slice)* | 2 x 1 | 1 x 1 | 2 x 1 | 2 x 1 | 2 x 1 | 1 x 1 | 2 x 1 |
| *NEX* | 1 | 1 | 0.7 | 1 | 0.7 | 1 | 1 |
| *BW (Hz/pixel)* | 70 | 488 | 325 | 122 | 325 | 488 | 70 |
| *TA (min:sec)* | 3:05 | 2:66 | 3:12 | 3:55 | 3:44 | 3:14 | 3:17 |

FSE fast spin echo; FS fat suppressed; PD proton density; SPGR spoiled gradient recalled echo; IDEAL-IQ iterative decomposition of water and fat with echo asymmetry and least-squares estimation; Uni unilateral; Bi bilateral; TR repetition time; TE echo time; FA flip angle; ETL echo train length; Acq acquired; Recon reconstructed; FOV field of view; Acc acceleration factor; NEX number of excitations; BW bandwidth; TA acquisition time.

##### Spine-to-Knee Protocol

###### 1. Axial IDEAL-IQ

An axial 3D iterative decomposition of water and fat with echo asymmetry and least-squares estimation (IDEAL-IQ) sequence was acquired for quantitative assessment of tissue fat composition in the lumbosacral spine. The sequence provides water- and fat-separated images for generation of fat fraction maps, enabling characterization of fat infiltration within the paraspinal musculature and assessment of bone marrow and intervertebral disc composition.

###### 2. Sagittal T_2_-weighted Cube with FS

A sagittal 3D T_2_-weighted Cube sequence with fat saturation was acquired for comprehensive evaluation of lumbosacral spine anatomy and fluid-sensitive pathology. The sequence provides high-resolution imaging for multiplanar assessment of the vertebral bodies, intervertebral discs, spinal canal, and surrounding soft tissues. The T_2_-weighted contrast provides sensitivity to disc degeneration, annular abnormalities, edema-like bone marrow changes, and other fluid-sensitive spinal pathology.

Acquisition parameters for lumbosacral spine MRI are provided in **Table 3**, and representative images are shown in **Figure 10**.

**Figure 10.**
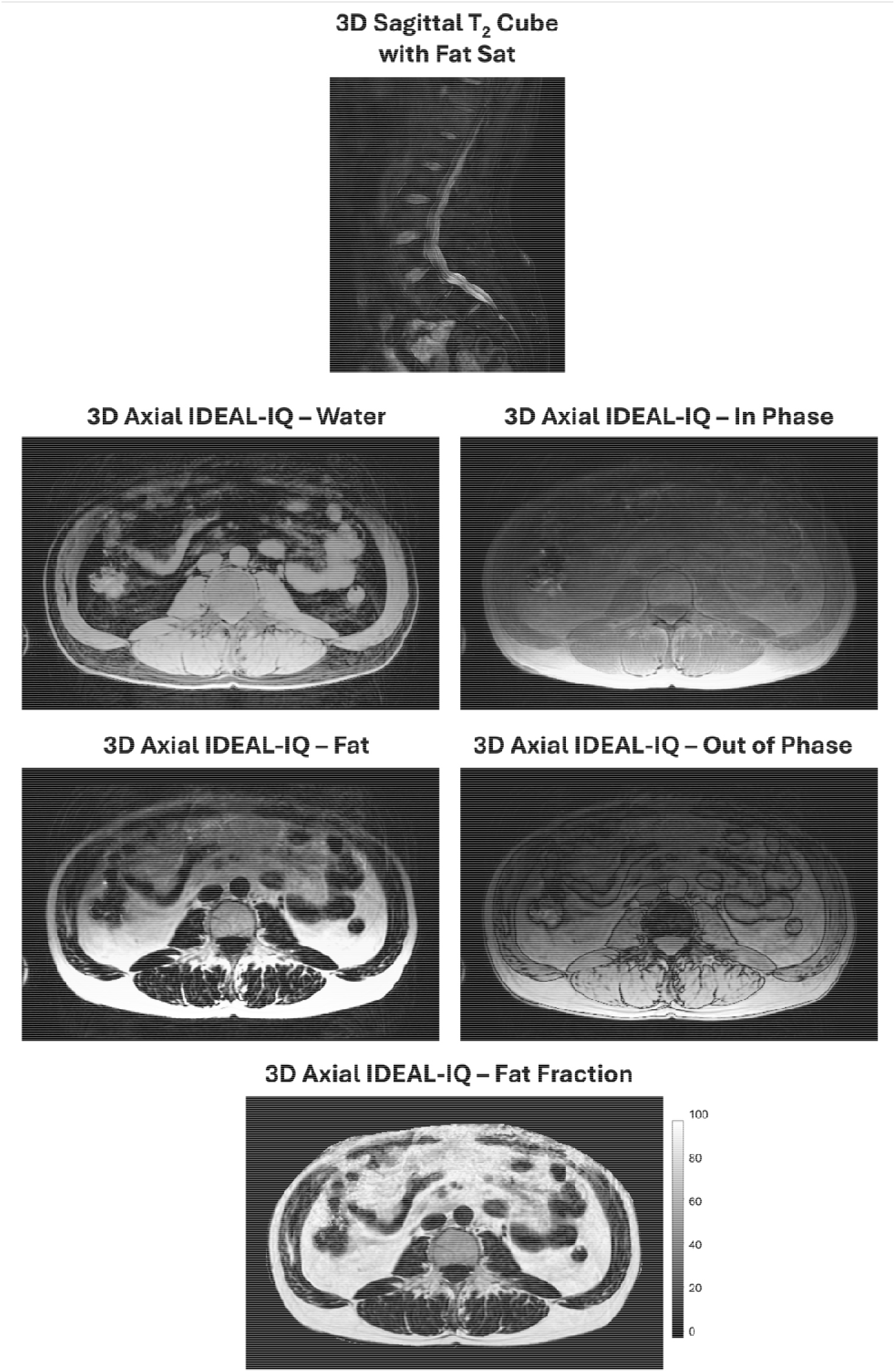
Representative lumbosacral spine MRI images, including axial IDEAL-IQ fat-water imagaes and sagittal T_2_-weighted Cube images with fat saturation. IDEAL-IQ images demonstrate quantitative assessment of fat composition within the paraspinal muscles, vertebral bone marrow, and intervertebral discs, while T_2_-weighted images provide fluid-sensitive visualization of the vertebral bodies, intervertebral discs, spinal canal, and surrounding soft tissues.

###### 1. Bilateral axial T_2_-weighted Cube with FS

A bilateral axial 3D T_2_-weighted Cube sequence with fat saturation was acquired for high-resolution, fluid-sensitive assessment of the hips and surrounding soft tissues. The sequence provides multiplanar visualization of the hip joints, including the femoral heads and acetabula, articular cartilage, labral region, tendons, muscles, and periarticular soft tissues. The fluid-sensitive contrast is particularly useful for detecting joint effusion, bone marrow lesions and edema-like changes, tendinopathy, muscle abnormalities, and other periarticular pathology.

###### 2. Bilateral coronal PD-weighted frFSE with FS

A bilateral coronal 2D PD-weighted frFSE sequence with fat saturation was acquired for comprehensive anatomical evaluation of the bilateral hip joints. The sequence provides high-resolution assessment of the articular cartilage, acetabular labral region, femoral head and neck, periarticular tendons and muscles, and surrounding soft tissues. The fat-suppressed PD contrast provides sensitivity to structural abnormalities, joint fluid, bone marrow lesions, and soft-tissue pathology while enabling direct comparison between hips.

Acquisition parameters for hip MRI are provided in **Table 3**, and representative images are shown in **Figure 11**.

**Figure 11.**
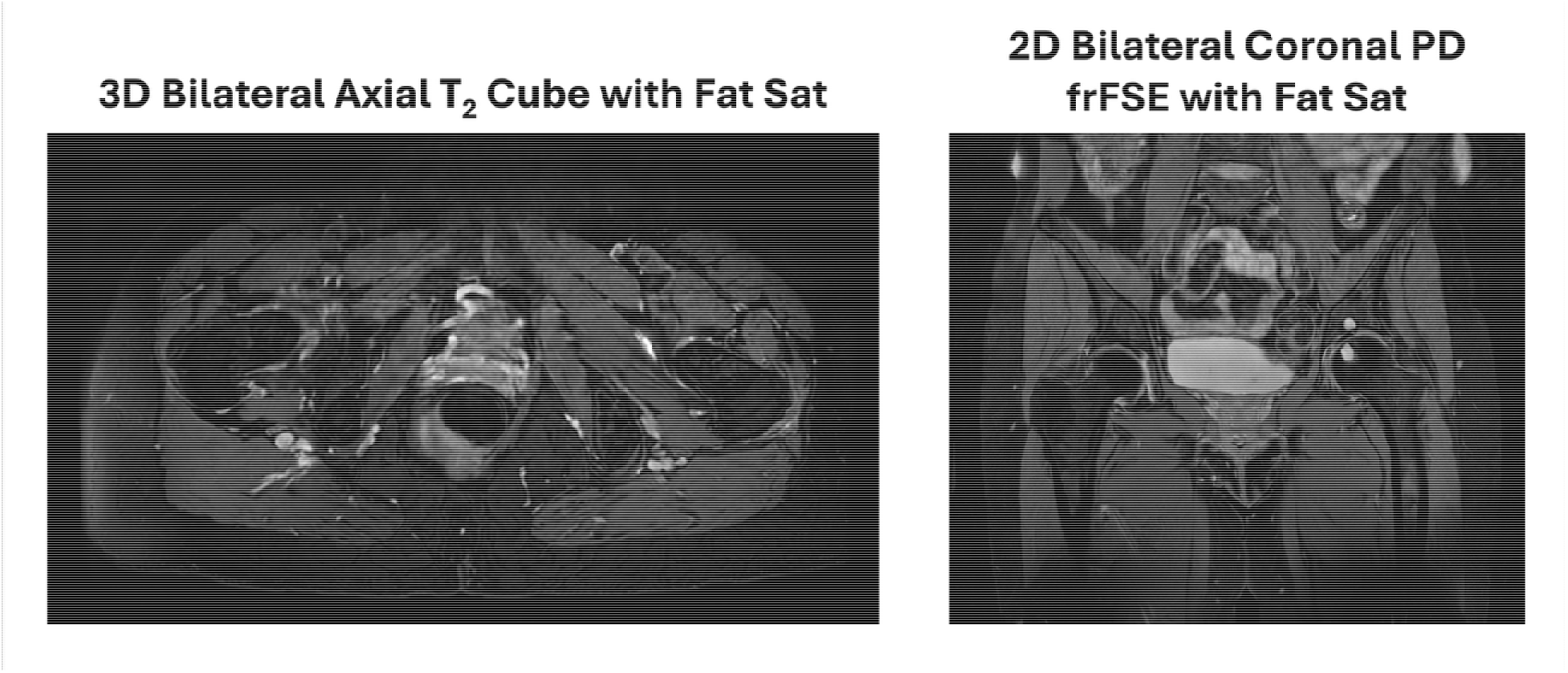
Representative bilateral hip MRI images from axial T_2_-weighted Cube with fat saturation and coronal PD-weighted frFSE with fat saturation sequences. The images demonstrate visualization of the femoral heads and acetabula, articular cartilage, labral region, bone marrow, periarticular tendons and muscles, and surrounding soft tissues, with fluid-sensitive sequences providing sensitivity to joint effusion, edema-like bone marrow abnormalities, and other periarticular pathology.

###### 1. Bilateral axial IDEAL-IQ

A bilateral axial 3D IDEAL-IQ sequence was acquired for quantitative assessment of thigh muscle composition. The sequence provides water- and fat-separated images for generation of fat fraction maps, enabling quantification of intramuscular fat infiltration across the major thigh muscle groups (hamstrings, quadriceps, and adductors). These measurements provide a quantitative marker of muscle composition and can be used to characterize regional differences in muscle quality.

###### 2. Bilateral axial SPGR

A bilateral axial 3D SPGR sequence was acquired to provide high-resolution anatomical imaging of the bilateral thighs. The sequence enables detailed characterization of muscle morphology and surrounding soft-tissue anatomy and provides a structural framework for potential future finite element analysis (FEA), including modeling of bone strain and geometry.

Acquisition parameters for thigh MRI are provided in **Table 3**, and representative images are shown in **Figure 12**.

**Figure 12.**
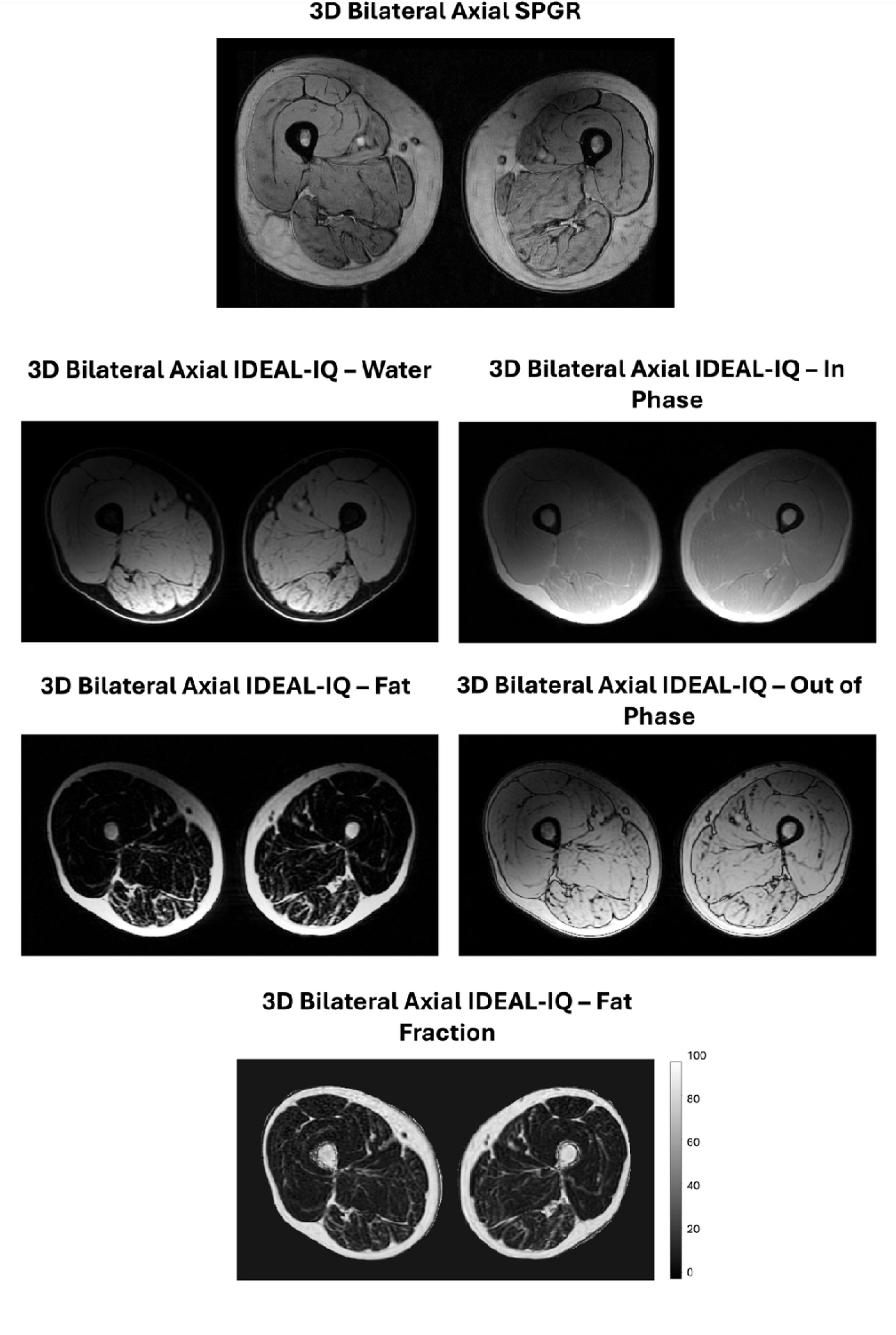
Representative bilateral axial thigh MRI images, including IDEAL-IQ fat-water images and 3D SPGR anatomical images. IDEAL-IQ images allow quantitative assessment of intramuscular fat infiltration within the quadriceps, hamstrings, and adductor muscle groups. SPGR images provide high-resolution anatomical visualization of muscle morphology and surrounding soft tissues and provide anatomical input for potential future finite element modeling of bone geometry and strain.

###### Longitudinal Follow-Up Scan MRI Protocol

At the 1-year and 2-year follow-up time points, a standardized subset of the baseline MRI protocol was repeated to enable longitudinal assessment of structural and quantitative changes. Knee MRI included sagittal T_2_-weighted frFSE with fat saturation, coronal PD-weighted FSE with fat saturation, axial PD-weighted frFSE with fat saturation, coronal T_1_-weighted FSE, sagittal T_2_/PD-weighted Cube with fat saturation, sagittal qDESS, sagittal B_1_ mapping, and sagittal four-echo UTE for quantitative T_2_/T_2_* mapping. Thigh MRI included axial IDEAL-IQ for assessment of muscle fat fraction. Lumbosacral spine MRI included sagittal T_2_-weighted Cube with fat saturation, and hip MRI included coronal 2D PD-weighted imaging with fat saturation. The repeated acquisitions were selected to provide longitudinal measures of joint structure, cartilage composition, meniscal and ligamentous properties, muscle composition, and spine and hip pathology while maintaining consistency with the baseline imaging protocol.

### 3. PET-MRI Image Analysis

#### 3.1 MRI-Based Segmentation

##### Knee Tissues

An open-source, automated image processing pipeline, OpenMSK[32], was used for tissue segmentation and quantitative morphometric analysis. OpenMSK is a modular deep-learning-based framework that accommodates multiple MRI acquisition protocols while providing standardized tissue segmentations and regional analyses. For this study, bilateral qDESS MRI scans were used to automatically segment the knee bones (femur, tibia, and patella), cartilage (femoral cartilage, medial and lateral tibial cartilage, and patellar cartilage), and menisci (medial and lateral).

Cartilage segmentations were further subdivided into anatomically meaningful regions for quantitative analysis, including five femoral subregions (anterior, medial central, medial posterior, lateral central, and lateral posterior), resulting in eight total regions (combined with two tibial and one patellar region). Each cartilage region was additionally divided into deep and superficial layers using a normalized thickness-based approach to enable depth-dependent analysis.

Subchondral bone regions were generated directly from the whole-bone and cartilage segmentations for PET analysis. For each bone, subchondral bone was defined as all bone voxels located within 3 mm of the overlying cartilage. This distance was selected to account for the limited spatial resolution of clinical [^18^F]NaF PET (intrinsic spatial resolution approximately 4–6 mm and our system’s reconstructed voxel size 1.3 x 1.3 x 2.78 mm^3^) while capturing tracer spill-in from the bone-cartilage interface. Subchondral bone regions were subdivided into the same anatomical regions as the corresponding eight cartilage to enable direct regional comparisons between PET and MRI biomarkers.

##### Thigh, Pelvic and Paraspinal Muscles and Bones

Muscle anatomy was segmented using the MuscleMap toolbox[33], which provides an automated segmentation framework to generate subject-specific masks of the thigh, pelvic, and paraspinal muscles, and corresponding osseous structures. Muscle segmentations included the major thigh, pelvic, and paraspinal muscle groups, while bone segmentations included the pelvis, femur, and tibia. The resulting segmentations were used to quantify muscle morphology and composition and to define anatomically resolved regions of interest for quantitative MRI analyses, including muscle fat fraction.

##### Spine

The TotalSpineSeg[34] automated segmentation framework, a part of the Spinal Cord Toolbox, was used to segment the lumbosacral spine, including individual vertebral bodies and intervertebral discs. The resulting segmentations provided anatomically defined regions of interest for quantitative analysis of spinal bone and disc imaging biomarkers and enabled assessment of tissue-specific changes across the spine.

#### 3.2 MRI Analysis

##### Cartilage T_2_ Relaxometry

Quantitative cartilage T_2_ maps were generated from the bilateral qDESS acquisitions using the two acquired echoes from the below equation and as described described [30]:

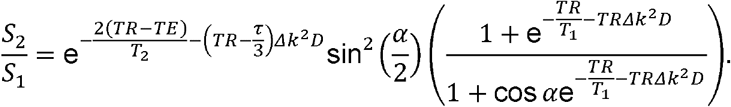

Where TR and TE represent the repetition and the first echo time, respectively, *α* is the flip angle, D is the diffusivity, and the dephasing per unit length induced by the unbalanced gradient is denoted by *Δk=γGτ* where G and *τ* are the spoiler amplitude and duration, respectively, and *γ* is the gyromagnetic ratio. A T_1_ relaxation time of 1.2 seconds and D = 1.25×10^−9^m^2^/s for cartilage is assumed in the model. Acquired B_1_ maps were used to adjust *α* in the model on a pixel-by-pixel basis[35]. The resulting maps were used to assess regional cartilage composition. All T_2_ data analysis was performed using OpenMSK[32].

##### Meniscus and Ligament UTE-T_2_* Relaxometry

Quantitative T_2_* maps were generated from the four-echo UTE acquisitions using a mono-exponential decay model with linear least-squares fitting. Voxel-wise T_2_* values were estimated from the signal decay across the four echo times and used to assess short-T_2_ tissues, including the menisci, tendons, and ligaments.

##### Muscle Fat-Water Composition

Quantitative fat-fraction maps were generated from the IDEAL-IQ acquisitions using the water- and fat-separated images[36]. Fat fraction was calculated voxel-wise as the ratio of the fat signal to the combined fat and water signal, providing a quantitative measure of tissue fat composition.

#### 3.3 PET Analysis

PET image processing and pharmacokinetic analysis were performed using an in-house MATLAB pipeline consisting of automated arterial input function (AIF) extraction, motion correction, anatomical registration, region-of-interest analysis, and compartmental kinetic modeling (github.com/agoyal5/dynafknee).

For each dynamic PET acquisition, an image-derived AIF was obtained from the PET angiography images using automated vessel segmentation. Briefly, peak tracer activity was identified, background activity was removed, and vessel candidates were refined using area-under-the-curve and temporal gradient analyses. Mean tracer activity within the segmented arterial voxels was then calculated over time to generate the AIF. The 3-minute dynamic acquisition, acquired immediately prior to the second [^18^F]NaF injection, was used to measure residual tracer activity from the baseline scan, which was incorporated into the pharmacokinetic model and accounted for during estimation of the post-exercise tissue time-activity curves (TACs).

Dynamic PET TAC images were rigidly co-registered to a common reference frame (same time as the qDESS MRI acquisition) using independent registrations for the left and right knees to minimize motion throughout each dynamic acquisition. qDESS-derived segmentations were resampled to PET space and used to extract regional TACs from each anatomical region.

Regional TACs were analyzed using the Hawkins two-tissue three-compartment model[26] implemented in COMKAT[37]. The measured TACs and image-derived AIF were used to estimate bone perfusion (K_1_), tracer washout (k_2_), and fluoride incorporation into bone mineral (k_3_)[22].

The extraction fraction (ExFr), representing the fraction of delivered tracer irreversibly incorporated into bone, was calculated as:

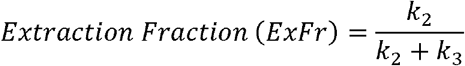

Net tracer influx (K_i_), the net rate of fluoride incorporation into bone and a quantitative measure of bone mineralization, was then calculated using non-linear regression, as:

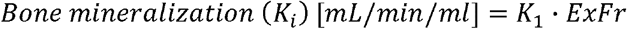

Standardized uptake values (SUV) were calculated from the static end-of-scan images. For each anatomical region, mean and maximum SUV, K_1_, K_i_, and ExFr were computed. SUV provides a semi-quantitative measure of tracer uptake, while K_1_, K_i_, and ExFr provide quantitative measures of tracer dynamics.

### 4. Video-Based Biomechanics

To assess whole-body biomechanics, we used OpenCap, an open-source, video-based motion analysis system[38]. Participants completed a standardized stair-climbing task consisting of eight flights of stairs, ascending and descending. Video recordings were acquired using two smartphones during the task and processed using OpenCap to estimate whole-body kinetics. Participants completed approximately 135 steps per side across the stair-climbing task, providing repeated measurements per trial.

### 5. Patient-Reported Outcomes

Participants reported average pain intensity separately for each knee using a 100-point visual analog scale (VAS), with 0 indicating no pain and 100 indicating the worst imaginable pain[39]. Participants also indicated the location of their knee pain on a standardized knee pain localization map[40], with pain categorized as medial, lateral, peripatellar, or infrapatellar.

Beginning with participant number 27 (additional spine-hip-thigh protocol), VAS pain assessments were additionally collected separately for both hips and the lower back to characterize pain at other anatomical sites.

The knee injury and osteoarthritis outcome score (KOOS)[41] was also collected for all participants. For participants with osteoarthritis or knee pain, KOOS scores were assessed for the more painful knee; for asymptomatic participants, the right knee was used as the index knee.

Physical activity was assessed using the international physical activity questionnaire (IPAQ)[42], a standardized questionnaire assessing habitual physical activity.

## Data Availability

All data produced in the present study will be shared publicly soon; in the meantime, we can make data available upon reasonable request to the authors.

## Acknowledgements

We would like to thank Dawn Holley, Andrew Dreisbach, Kim Halbert, and Dr. Mehdi Khalighi for their support with PET-MR imaging.

